# Synthesising Interictal Epileptiform Discharges With Generative Adversarial Network

**DOI:** 10.1101/2025.09.06.25335239

**Authors:** Duong Nhu, Mubeen Janmohamed, Patrick Kwan, Piero Perucca, Terrence J. O’Brien, Amanda K. Gilligan, Zonguan Ge, Chang Wei Tan, Levin Kuhlmann

## Abstract

Interictal epileptiform discharges (IEDs) are reliable biomarkers in electroencephalograms for epilepsy. To automate IED detection, deep learning (DL) has been applied to treat EEG signals as time-series data. However, collecting sufficient IEDs for training DL models is expensive; public and private datasets are often overwhelmed with background activities. To address this issue, generative adversarial networks (GANs) have been used to synthesise IEDs. Previous works using GANs achieved some improvements, but the quality metrics of synthetic samples were not contextualised, resulting in suboptimal model tuning. In this work, we proposed to use density and coverage to evaluate synthetic data and to tune a GAN model. We also utilised soft dynamic time warping (soft-DTW), a differentiable version of DTW, as the reconstruction loss to learn temporal alignments of the IED’s waveforms and improve the diversity of synthetic samples. Our proposed approach achieved a 10% improvement in the AUPRC score on the Temple University Events (TUEV) dataset by combining synthetic and real data, demonstrating the potential of this method to enhance the accuracy of IED detection.

## 1 Introduction

The diagnosis and monitoring of epilepsy often require the assessment of interictal epileptiform discharges (IEDs) through scalp EEG recordings. IEDs are characterised by intermittent patterns of spikes, sharp waves, or bursts of fast activities that stand out from background activities. However, the process of analysing EEG to identify IEDs is time-consuming and arduous, sometimes taking several hours. To address this challenge, deep learning (DL) has been applied to automate IED detection [20, 19, 12, 3]. Most DL models consider EEG signals as time-series data and use temporal convolutional and recurrent networks to distinguish IED samples from background activities. Nonetheless, the slow and costly IED labelling process remains a bottleneck to achieving high-performance IED detectors (i.e. AUPRC, F1, etc.), given that IEDs are often outnumbered by background activities in existing datasets [20], causing the model to be biased towards negative samples. Oversampling IEDs imbalances the data but does not introduce new data and might result in overfitting. Synthetic minority oversampling technique (SMOTE) [1] or adding random noise can bring in more new data; however, these samples might not represent the real data and contain unnecessary noise. A more reliable solution is to use generative adversarial networks (GANs) to synthesise IED for augmentation [7, 6, 22].

Existing works on GAN for IED synthesis often measure the quality of the generator by fitting a classifier on the generated samples, which is a lengthy process. In image classification, the Frechet Inception Distance (FID) [9] is often used to measure the quality of the synthetic samples. As the majority of the existing works in automated IED detection used private datasets without benchmarking on public datasets [20], choosing a state-of-the-art architecture to calculate a similar score to FID is ambiguous. Furthermore, FID does not measure fidelity and diversity [18] that are crucial for IED synthesis evaluation. Assessment via visualisations is not feasible as it requires human experts. Therefore, there is a need for more contextualised and reliable quantitative metrics to quickly measure the quality of synthetic IEDs and tune hyperparameters in GANs.

In IED detection, EEG signals are split into small windows of 1 or 2 seconds [20], but a single IED lasts between 20 and 200 milliseconds [15] and can occur anywhere in a window. Hence, element-wise comparisons of corresponding timesteps or cross-entropy loss do not take the time shift problem into account. Dynamic time warping (DTW) is widely used in time-series tasks as it can find the optimal alignment between 2 time-series whose data points are not synchronised [17]. As such, DTW has also been used in IED detection [25, 11]. However, DTW is not differentiable[2], which limits its usefulness. Soft-DTW [2] has been proposed to solve this problem by introducing a smoothing parameter. This property makes soft-DTW a suitable loss for a GAN network, which might improve the diversity of synthetic samples.

In this work, we used density and coverage, as described in Naeem et al. (2020) [18], to measure the quality of synthetic IEDs and to tune a modified AlphaGAN [23] architecture. We also experimented with our architecture with and without soft-DTW as the reconstruction loss. Finally, we used the synthetic IEDs as augmented training samples for a classifier. Our results showed that our approach could speed up the tuning process and produce reliable results.

## 2 Method

### 2.1 AlphaGAN

The AlphaGAN [23] framework is a successful solution to prevent mode collapse in generative models by combining a variational autoencoder and a GAN. We modified the AlphaGAN architecture to generate IED samples. AlphaGAN comprises four components: *generator, discriminator, encoder*, and *code discriminator*. These components are illustrated in Figure 1. The *encoder* learns the embedding 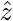 of the real samples, whereas the *generator* decodes the latent space and reconstructs the real samples from the embedding outputs generated by the encoder. The *generator* is optimised to maximise the discriminator loss and minimise the reconstruction loss. The *code discriminator* distinguishes the embedding of real samples as fake and the embedding of fake samples as real. On the other hand, the *discriminator* labels the reconstructed and generated samples as fake and the real samples as real. For a detailed description of the training process, we recommend referring to the original AlphaGAN work [23].

**Fig. 1.**
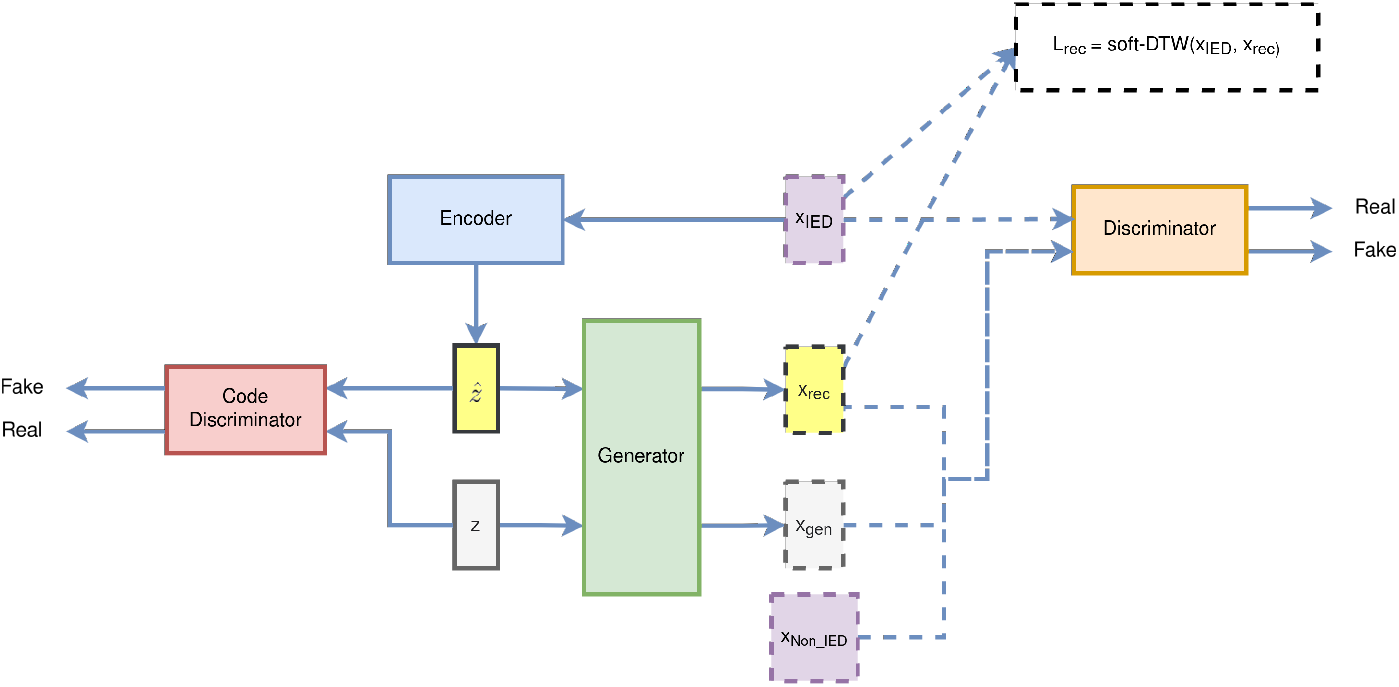
Components of our AlphaGAN architecture. The dashed lines highlight our modifications. The generator only generates IED samples. The soft-DTW loss is used as a reconstruction loss. Non-IED samples are passed to the discriminator as fake data.

We evaluated the AlphaGAN with mean absolute percentage error distance and with soft-DTW (described below) to compare the similarity between real samples and reconstructed samples. In addition, we only pass the IED samples to the encoder. The non-IED samples were treated as fake samples when updating the discriminator. The reconstruction loss was weighted by a *λ* coefficient. The latent space, *z*, of fake samples was sampled from a uniform sphere distribution.

### 2.2 Choice of reconstruction loss

Element-wise comparisons of timesteps may miss important information that positions of IEDs vary across windows, but their morphisms are similar. To accurately reconstruct the time-variance of IEDs, we employed the Soft Dynamic Time Warping (Soft-DTW) algorithm [2] as a reconstruction loss, in addition to mean absolute percentage error (MAPE). Soft-DTW is a differentiable and smooth version of DTW that measures the similarity between two time-series, considering time shifts. Previous studies have demonstrated the efficacy of DTW in clustering IED windows [25, 11]. Unlike DTW, Soft-DTW provides a continuous gradient, which can be computed using the chain rule, making it ideal for backpropagation in deep learning models.

Soft-DTW introduces a smoothing parameter *γ* to control the smoothness of the warping function, and is defined as follows,

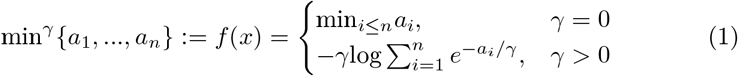

When *γ* = 0, soft-DTW becomes the original DTW. If *γ >* 0, soft-DTW can be explicitly differentiated. Small values of *γ* were shown to have better performance [2]. In our experiments, we used soft-DTW with MAPE and *γ* = 0.001.

### 2.3 Architecture choices and baseline

InceptionTime has been shown to have great performance in IED detection [19]. We used InceptionTime with the same configurations as in Nhu et al. (2022) [19] as the base classifier, and InceptionTime with a few modifications as the architectures for the *encoder, generator*, and *discriminator*. We used the same parameters for the InceptionTime in all components. For the encoder, we replaced the global average layer by flattening all dimensions and passing them to a linear operation of 64 units. This was the same as in the discriminator, which was followed by a binary classification layer. The last layer of the generator was a convolution with a stride of 1, mapping each timestep to the same dimensions as in the real data. The discriminator and generator did not use batch normalisation. Instead, we used spectral normalisation [16], which was shown to be effective even without Wasserstein loss. The encoder and the generator used ReLU. The discriminator used LeakyReLU with *α* = 0.2. We provided more details of the settings in Table S1, S2, and S3 in the supplement.

The code discriminator was a 3-layer self-normalisation network (SNN) [13]. Each layer had 750 units, SeLU as the activation function, and was followed by an alpha dropout layer with a rate of 0.05.

### 2.4 Quality measurement of generated samples by the generator

Diversity measurement of GAN-generated samples has been extensively explored in prior research [24, 14, 18]. Sajjadi et al. (2018) [24] introduced *precision* and *recall* as indicators of the quality of generated images, where *precision* measures the realism of generated images and *recall* denotes the degree to which the generator covers the training data. Kynkäänniemi et al. (2019) [14] proposed *improved precision* and *recall* metrics by using k-nearest neighbour distances to estimate the probability density functions. However, it is noted that these metrics may be sensitive to outliers [18].

To address the issue of the above metrics, Naeem et al. (2020) [18] proposed *diversity* and *coverage. Density* improves upon *precision* by counting the number of real sample neighbourhood spheres containing fake samples. *Coverage* improves upon *recall* by measuring the fraction of real samples whose neighbourhood contains at least 1 fake sample. By incorporating these metrics, the evaluation of GAN models becomes more robust and less affected by outliers.

We denote feature vectors of real and generated samples as *ϕ*_*r*_ and *ϕ*_*g*_, and the respective sets as *Φ*_*r*_ and *Φ*_*g*_. We define a binary function *f* (*ϕ, Φ*),

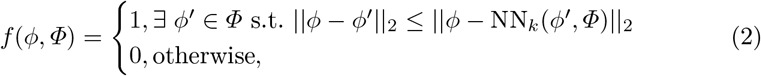

where NN_*k*_(*ϕ*^*′*^, *Φ*) returns the k*th* nearest feature vector of *ϕ*^*′*^ from set *Φ. Density* and *coverage* are defined as follows,

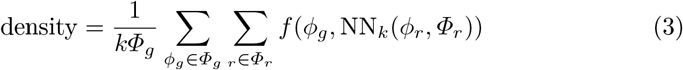

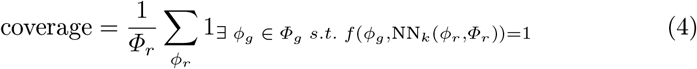

where 1_(.)_ is the indicator function.

We used the pre-trained encoder to generate the embeddings for the real and synthetic IED samples. Naeem et al. (2020) [18] suggested using the random-initialised network to generate the embeddings; however, we found a pre-trained encoder more reliable, as the results were consistent. The number of neighbours was set to 5. The number of synthetic and real samples was the same.

## 3 Datasets

### 3.1 Public dataset - Temple University Events (TUEV)

The Temple University Hospital (TUH) corpus [21] is currently the most extensive publicly available EEG corpus related to epilepsy. It comprises multiple datasets designed for different purposes, such as seizure detection, artifact detection, and epileptiform discharge detection. For this study, we utilised the Temple University Events (TUEV) dataset [8], which was specifically curated for identifying IEDs. The TUEV dataset included 3 types of events: spike and sharp wave (SPSW), periodic lateralized epileptiform discharges (PLEDs), generalised periodic epileptiform discharges (GPEDs), and non-IEDs (artifacts, eye movements, and background). Each channel in the EEG epoch was labelled with a corresponding event, even if the event was present across all channels. As such, we treated these IEDS as single-channel time-series. Although there is an ongoing debate on whether GPEDs and PLEDs should be classified as IEDs or not [5], we chose to exclude them from our work to avoid any ambiguity. Since SPSW can originate from patients with generalised or focal epilepsy, we treated SPSW as single-channel time-series. The numbers of samples of these events are given in Table 1.

**Table 1.** Number of annotations in TUEV train & evaluation sets.

| Annotations | Train set | Evaluation set |
| --- | --- | --- |
| SPSW | 645 | 567 |
| GPED | 11,229 | 4,672 |
| PLED | 5,965 | 1,998 |
| Non-IED | 65, 109 | 22,134 |

**Table 2.** Density and coverage scores for the best *λ*_*rec*_ with and without soft-DTW for each dataset.

| Dataset | Density | Coverage | $\lambda_{rec}$ | soft-DTW |
| --- | --- | --- | --- | --- |
| SPSW - TUEV | 2.81 | 0.49 | 50 | yes |
| Alfred | 1.01 | 0.03 | 5 | no |

### 3.2 Private datasets

We collected routine EEG recordings from patients with idiopathic generalised epilepsy (IEG) at two national public hospitals, Alfred Health and Royal Melbourne Hospital (RMH). We also gathered normal EEG recordings that did not show any IEDs. The datasets collected from these two hospitals contained only SPSW and were annotated epoch-wise. This means that any multichannel epoch that had SPSW was labelled as IED. The annotation of the recordings was conducted by three board-certified neurologists who marked the beginning of the annotation at 100-200 milliseconds before the first visible spike or rhythm change, and the end of the annotation at the last slow-wave’s end. The combined EEG recordings from both hospitals yielded a total of 1,155 IEDs and 1,608 IEDs in the Alfred and RMH datasets, respectively. In this work, we trained our AlphaGAN on Alfred, fitted a classifier on augmented data and tested the generalizability on RMH. Alfred Health Ethics Committee approved this study (Project No: 745/19).

## 4 Experimental setup

### 4.1 Univariate and multivariate tasks

Based on the characteristics of different IED categories and how each dataset was annotated, we divided our experiments into 2 tasks: ***univariate*** and ***multivariate***. For the univariate task, we aimed to improve SPSW detection in the TUEV dataset. PLEDs are repetitive SPSW and are limited to a focal brain area [8]. GPEDs are similar to PLEDs but are observed across both hemispheres [8]. As such, it is intuitive to detect PLEDs and GPEDs by using multichannel epochs. We used these for the multivariate task. The Alfred and RMH datasets, which were annotated epoch-wise and only contained SPSW, were likewise used for the multivariate task.

### 4.2 Preprocessing

To ensure the accuracy of our results, we curated our private datasets, specifically using only normal EEG recordings for the negative class to prevent any missed IEDs by the neurologists. We standardised our dataset by excluding auricular electrodes, M1 and M2, as they were not present in several EEG recordings, leaving us with 19 electrodes for each recording. To ensure consistency across all datasets, we applied the Temporal Central Parasagittal (TCP) montage from the TUEV dataset to all recordings. Additionally, we resampled all EEG recordings to 256 Hz using polyphase filtering and applied bandpass filters ranging from 0.5-49 Hz to eliminate high and slow-frequency artifacts and power-line noises.

### 4.3 Data augmentation with synthetic data

The study findings revealed that the synthetic data generated was not without imperfections, as later demonstrated. Aiming to avoid the introduction of unwanted noise through the generation of an excessive amount of synthetic data for the purpose of balancing the dataset, alternative approaches were explored. Specifically, different ratios of both real and synthetic IEDs and background samples were examined, with values of 0.5 and 0.3 being tested. Additionally, label smoothing was applied to prevent the classifier from learning from noise, with smoothing factors of 0.1 and 0.2 being evaluated. Label smoothing was only applied to synthetic IEDs, while the labels of real IEDs remained unaltered.

### 4.4 Training configurations

We used Adam optimiser for all components of AlphaGAN but with *β*_1_ = 0.5 and *β*_2_ = 0.9. The learning rate of the generator was 0.0001, and that of the other components was 0.0005. For the code discriminator, we set Adam’s epsilon to 0.1. We trained AlphaGAN for 100 epochs, then if the validation AUPRC of the discriminator did not improve after 15 epochs, the training process was stopped.

## 5 Results

### 5.1 Quality of generated samples

Table 3 demonstrates density and coverage for the best *λ*_*rec*_ for each dataset. While for TUEV, *λ*_*rec*_ = 50 with soft-DTW achieved the highest performance, *λ*_*rec*_ = 5 with only MAPE had the best performance in Alfred. The generator trained on Alfred had a density of 1.01, which means it could generate realistic samples; however, its low coverage indicates mode collapse. In contrast, the generator trained on TUEV had high density and coverage, whose samples are illustrated in Figure S1 in the supplement. These results suggested that our AlphaGAN architecture struggled to generate multichannel IED samples but could successfully generate single-channel IED samples. Table 3 showed the density and coverage scores for the best *λ*_*rec*_ for each dataset. We also compared results with soft-DTW to simple MAPE. The full lists of scores for each configuration are provided in Table S1 and S2 in the supplement. AlphaGAN with soft-DTW models outperformed simple MAPE in most tasks, except for generating PLED.

**Table 3.** Density and coverage scores for the best *λ*_*rec*_ in TUEV and A datasets.

| Dataset | $\lambda_{rec}$ | Density | Coverage | soft-DTW |
| --- | --- | --- | --- | --- |
| TUEV | 50 | 2.81 | 0.49 | yes |
| Alfred | 5 | 1.01 | 0.03 | no |

### 5.2 IED detection with augmented data

In this experiment, we used the synthetic data for augmentation and combined it with the real data to train a classifier. We also trained a baseline classifier in which IEDs were oversampled to balance the dataset, and no data augmentation was used. For threshold-based metrics, we chose the probability threshold to maximise the F1 score in the validation set.

Table 4 shows the classification performance on the TUEV dataset. The classifier fitted on real and synthetic samples with label smoothing of 0.2 and a ratio of 0.5 had the best performance with an AUPRC of 0.8, which is a 10% increase compared to the baseline. We also noted that our work was better than the InceptionTime with log transformation by Duong Nhu et al. (2022) [19], which was trained on only TUEV data and used mixup [10] for data augmentation.

**Table 4.** Performance of baseline and augmented models on TUEV’s evaluation set.

| Model | Ratio | Label<br>smoothing | Sensitivity | Precision | F1 | AUPRC |
| --- | --- | --- | --- | --- | --- | --- |
| Baseline | N/A | N/A | 0.59 | 0.79 | 0.61 | 0.60 |
| Augmented | 0.5 | 0.1 | 0.92 | 0.30 | 0.45 | 0.70 |
|  | 0.3 | 0.1 | 0.88 | 0.35 | 0.50 | 0.68 |
|  | 0.5 | 0.2 | 0.90 | 0.50 | 0.63 | **0.80** |
|  | 0.3 | 0.2 | 0.93 | 0.28 | 0.43 | 0.66 |
| InceptionTime - Log [19] | N/A | N/A | 0.61 | 0.61 | 0.61 | 0.70 |

Table 5 provides the results of the classification performance on the Alfred. Despite not having good coverage, the model trained on synthetic and real data showed a slight improvement in AUPRC compared to the baseline. We also tested these models on RMH and observed an increase in AUPRC from 0.61 to 0.72. These results indicate that we can use density and coverage to further tune the AlphaGAN and InceptionTime architectures to achieve better performance.

**Table 5.** Results of baseline and augmented classifiers trained on Alfred. We also tested these models on the RMH.

| Dataset | Model | Ratio | Label<br>smoothing | Sensitivity | Precision | F1 | AUPRC |
| --- | --- | --- | --- | --- | --- | --- | --- |
| Alfred | Baseline | N/A | N/A | 0.40 | 0.74 | 0.52 | 0.56 |
|  | Augmented | 0.5 | 0.1 | 0.44 | 0.75 | 0.55 | 0.58 |
|  |  | 0.3 | 0.1 | 0.45 | 0.75 | 0.56 | **0.59** |
|  |  | 0.5 | 0.2 | 0.43 | 0.73 | 0.54 | **0.59** |
|  |  | 0.3 | 0.2 | 0.43 | 0.73 | 0.54 | 0.57 |
| RMH | Baseline | N/A | N/A | 0.60 | 0.59 | 0.59 | 0.61 |
|  | Augmented | 0.3 | 0.1 | 0.64 | 0.76 | 0.69 | 0.72 |

## 6 Discussions

By incorporating soft-DTW reconstruction loss into AlphaGAN, we were able to generate high-quality IED samples. These were shown via higher density and coverage scores than the MAPE loss, except for synthesising GPLED. Using these metrics to choose the best set of synthetic samples as augmented data, we managed to improve the IED detection performance. This indicates that coverage and density are suitable metrics to measure the fidelity and diversity of IEDs and time-series data. To assess the quality of our synthetic samples and the optimality of our AlphaGAN model, we utilised density and coverage measures. Our approach produced realistic and diverse single-channel IED samples when we applied soft-DTW. However, generating multichannel IED samples for generalised epilepsy in the Alfred posed a challenge. We found that combining synthetic samples with real data from TUEV enhanced classifier performance. Furthermore, we observed that the ratio of synthetic to real samples also had an impact on the performance. Our findings demonstrate that density and coverage metrics are effective for assessing the fidelity and diversity of synthetic time-series data. This approach can be applied to tune the InceptionTime architecture to generate better IEDs.

## 7 Conclusion

Our work demonstrated that density and coverage were suitable for assessing the quality of synthetic IEDs from GAN and could be used to tune hyperparameters. Our AlphaGAN architecture, combined with soft-DTW as a reconstruction loss function, could produce high-quality and diverse single-channel IED samples. However, generating multichannel samples was still a challenge. Additionally, by using these synthetic samples as augmented data, we have achieved an improvement in IED detection performance. Our methodology has the potential to extend to the synthesis of other types of physiological data where annotations are limited. Moreover, this can be applied to other time-series tasks beyond physiological research. For instance, our proposed architecture could be employed to synthesise realistic samples and address the imbalanced class issue in widely-used time-series classifiers such as the UCR archive [4].

## Data Availability

All data produced in the present study are available upon reasonable request to the authors.

## Appendix

**Table 6.** Metrics of synthetic samples from the Alfred dataset. AlphaGAN with *λ*_*rec*_ = 50 and soft-DTW had the best *F* 1_*DC*_ score.

| density | coverage | $F1_{DC}$ | $\lambda_{rec}$ | soft-DTW |
| --- | --- | --- | --- | --- |
| **149.40** | **0.73** | **0.85** | **50** | **Y** |
| 147.40 | 0.72 | 0.83 | 10 | N |
| 146.40 | 0.72 | 0.83 | 50 | N |
| 145.67 | 0.72 | 0.83 | 10 | Y |
| 114.80 | 0.56 | 0.65 | 5 | N |
| 90.19 | 0.44 | 0.51 | 5 | Y |
| 86.64 | 0.43 | 0.49 | 1 | N |
| 79.05 | 0.40 | 0.45 | 1 | Y |

**Table 7.** Metrics of synthetic GPLED and PLED samples from TUEV dataset. We calculated the average *F* 1_*DC*_ of GPED and PLED. AlphaGAN with *λ*_*rec*_ = 5 and soft-DTW had the best average *F* 1_*DC*_ score.

| Gped | | | Pled | | | | Avg $F1_{DC}$ |
| --- | --- | --- | --- | --- | --- | --- | --- |
| density | coverage | $\lambda_{rec}$ | density | coverage | $\lambda_{rec}$ | soft-DTW | |
| 224.90 | 0.49 | 5 | 345.05 | 0.46 | 5 | N | 0.63 |
| **223.60** | **0.49** | **5** | **355.93** | **0.47** | **5** | **Y** | **0.65** |
| 222.62 | 0.50 | 50 | 345.05 | 0.46 | 50 | N | 0.64 |
| 221.77 | 0.48 | 50 | 344.52 | 0.45 | 50 | Y | 0.63 |
| 221.76 | 0.48 | 10 | 343.22 | 0.44 | 10 | Y | 0.63 |
| 220.95 | 0.49 | 10 | 343.39 | 0.45 | 10 | N | 0.63 |
| 171.21 | 0.48 | 1 | 283.95 | 0.47 | 1 | N | 0.59 |
| 1.65 | 0.36 | 1 | 0.50 | 0.06 | 1 | Y | 0.01 |

**Table 8.** InceptionTime’s parameters.

| Hyperparameters | Values |
| --- | --- |
| Maximum kernel size | 32 |
| Number of filters | 64 |
| Number of convolutional blocks | 8 |
| Spatial dropout rate | 0.1 |
| Increase factor for the number of filters | 1 |

**Table 9.**
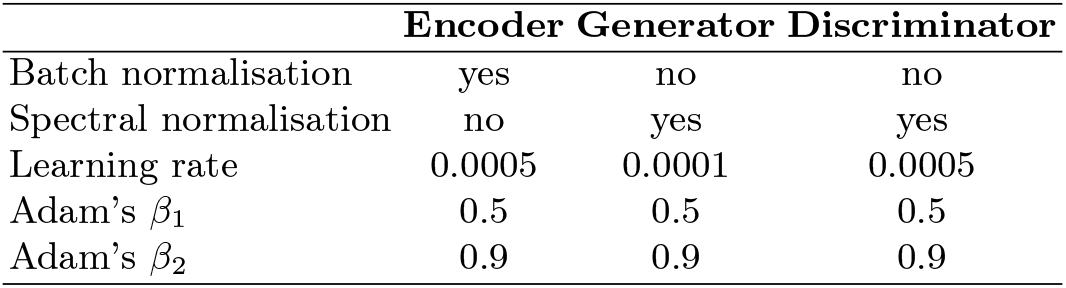
Settings of the encoder, generator and discriminator in our AlphaGAN architecture.

**Table 10.** Settings of the code discriminator of our AlphaGAN architecture. The code discriminator is a self-normalised network.

| Settings | Values |
| --- | --- |
| Number of layers | 2 |
| Alpha dropout rate | 0.1 |
| Number of hidden units | 750 |
| Learning rate | 0.0005 |
| Adam's $\beta_1$ | 0.5 |
| Adam's $\beta_2$ | 0.9 |
| Adam's $\epsilon$ | 0.1 |

**Fig. 2.**
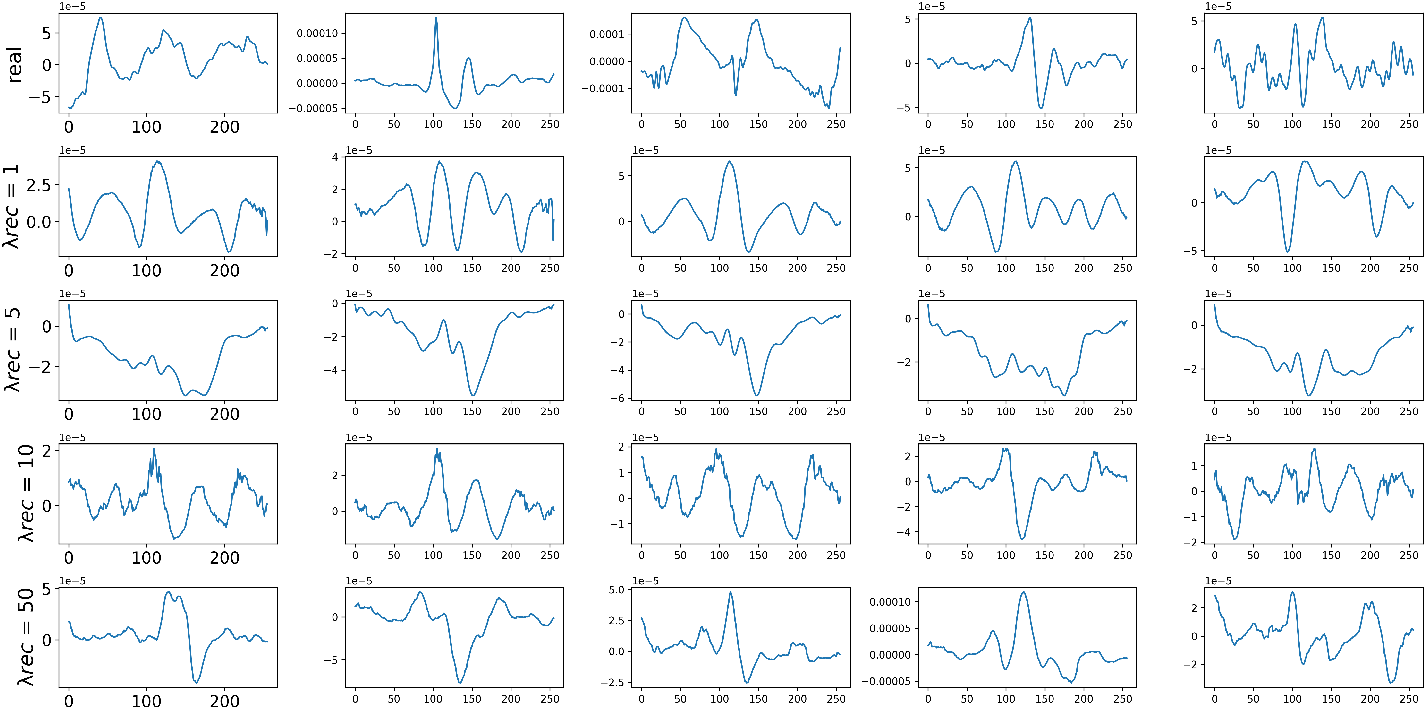
Synthetic SPSW from TUEV dataset generated by AlphaGAN. Each row has 5 figures corresponding to a *λ*_*rec*_. *λ*_*rec*_ = 50 had the best density and coverage.

